# Outdoor Scene Classrooms arrest myopia development: A school-based Randomized Clinical Trial

**DOI:** 10.1101/2025.03.10.25323549

**Authors:** Wei Pan, Longbo Wen, Xin Yi, Yongxiang Gong, Yang Zhe, Zhiwei Luo, Xiaoning Li, Guishuang Ying, Ian Morgan, Daniel Ian Flitcroft, Zhikuan Yang, Weizhong Lan

## Abstract

Reduced time outdoors is an important factor promoting the development of myopia. The spatial frequency spectra of man-made indoor and outdoor environments, which are impoverished in high spatial frequencies (HSF), may be a contributor to human myopia development. This randomized clinical trial aimed to investigate whether HSF enhancing classroom-- outdoor scene classroom (OSC) can arrest myopia development in children. Here we showed the OSC was protective in slowing down myopic shift among children who were not myopic at baseline. The one-year change of spherical equivalent was 0.22D less among baseline hyperopic children, and 0.18D less among baseline emmetropic children in the OSC group compare to traditional classroom group.

The protective effect of OSC is comparable to the time spent outdoors (Efficacy: OSC=37.1% vs. Outdoor=19.2%) and the 0.01% atropine (Efficacy: OSC=42.4% vs. 0.01% Atropine=30.9%). The study found that the OSC was protective in myopia prevention, especially in hyperopic children. Large implementation of OSC provides an alternative strategy to increased time outdoors in myopia prevention, and provides an approach that involves less disruption to school routines. Chinese Clinical Trial Registry identifier: ChiCTR2000040704.

## Introduction

Myopia, commonly known as short-sightedness, is a condition in which images of distant objected are focused in front of the retina, resulting in blurred distance vision. In addition to causing poor distance vision, myopia and particularly high myopia increases the risk of developing other conditions such as retinal detachment, myopic macular degeneration and a range of other ocular pathologies that cannot be corrected optically, and which may lead to severe visual impairment and blindness [1]. It has been predicted that 50% and 10% of global population will be myopic or be highly myopic by 2050 [2]. Obviously, the best way to avoid the complications associated with myopia and high myopia is to prevent the initial development of myopia. Reduced time spent outdoors, implying increased time indoors, is an important factor promoting the development of myopia, and increasing the amount of time that children spend outdoors has emerged as widespread preventive intervention [3].

More recently, Flitcroft et al. proposed that the spatial frequency spectra of man-made indoor and outdoor environments, which are impoverished in high spatial frequencies (HSF), may be a contributor to human myopia development [4]. These man-made environments share the property of lack of HSF components with visual environments created in experimental animal models of myopia, such as deprivation myopia and lens-induced myopia. In animals, it has been shown reductions in retinal image quality can cause stimulated myopia growth in many species [5–7]. In form deprivation myopia models, reduced retinal image quality can be achieved by covering animals’ eyes with diffusers of Bangerter foils [5,8]. The spatial frequency content of man-made indoor environments is similar to that achieved by placing a 0.4 Bangerter foil over a natural scene, which is able to induce myopia in species such as chicks and guinea pigs [5,9]. It is not known how much reduction in spatial frequency contents can act as myopiagenic stimuli in school aged children. However, even brief exposure to "normal" spatial frequencies significantly mitigates the effects of form deprivation myopia [7].

To investigate whether enhancing HSF components in indoor environments can arrest myopia development in children, we designed the outdoor scene classroom (OSC). The OSC was designed to resemble the natural outdoor environment by adorning the walls with wallpaper featuring natural images with high spatial frequency content. We hypothesized that students studying in the outdoor scene classroom would develop less myopia than those in traditional classrooms. This hypothesis was then tested in a two-arm randomized controlled trial.

## Methods

### Study Design, Setting, and Participants

The Outdoor Scene Classrooms Study (OSCS) is a two-arm, parallel, non-blinding, cluster randomized trial conducted in Lijiang, Yunnan, China. The study was designed to observe for two years from 01 Mar, 2021 to 01 Mar, 2023. Baseline visit was in March 2021 and the 1-year follow-up visit was in April 2022. Unfortunately, the trial was terminated in September 2022, before the 2-year follow-up visit, as the Lijiang Shiyan School had re-allocated students to a different campus due to administrative reasons. The current study reported results from baseline and 1-year follow-up visit.

A detailed study protocol and design was previously reported [10]. Full study protocol and statistical analysis plan can be viewed from the Supplementary materials. Briefly, randomization was achieved by cluster randomization, each individual class was treated as a cluster, 10 classes were randomized into intervention and control group with 1:1 ratio. Uniformly distributed random number range from 1 to 100 was generated for each class, and the 5 classes with largest random numbers were allocated to the intervention group (the OSC group), and the rest were randomized to the control group (the traditional classroom group). Random numbers were generated by our statistician, Lun Pan, using SPSS 24.0 software (IBM®) with seed number of 20201231. Due to the nature of the intervention, masking to participants was not possible, but the examiners and statistician remained blinded to the allocation during data collection and analysis processes.

Recruitment period took place from 01 Feb, 2021 to 01 Mar, 2021, and performed by Zhiwei Luo. As no blinding was applied to the participants, group allocation was told to the students, teachers, and school by Zhiwei Luo at baseline without any form of concealment.

All classes from Grade 3 in the Lijiang Shiyan school were included in this study. Students were excluded from the study if the student or their guardians refused permission to participate the study. Other exclusion criteria were: 1) unwilling or unable to administer cycloplegia (ie., including medication allergy, high intraocular pressure, and others); 2) strabismus or amblyopia; 3) using myopia control interventions (including but not limited to low-dose atropine, peripheral defocus spectacles, orthokeratology lens); 4) incomplete data; 5) systemic and/or ocular organic diseases that are relevant to myopia.

### Intervention

The intervention was the outdoor scene classroom, and the control was the traditional classroom. The outdoor scene classroom was designed to mimic the spatial frequency spectrum of natural environments. In the classroom, the interior design, including walls, ceilings, floors, and even students’ desks were covered with a specially designed wallpaper. The design consisted of images of natural scenes with green trees, blue skies, birds, etc. Images of natural world had been found to follow a relationship that the linear slope of log amplitude against log spatial frequency lies between -0.8 to -1.5 [4,11]. Flitcroft et al. had reported that the natural outdoor scene tended to have the slope around -1, and the indoor scene tended to be around -1.5 [4]. In the outdoor scene classroom, the spatial frequency contents of the design were required to have the slope higher than -1.15. In the traditional classroom, original white walls, ceilings, and plain floors, desks were retained. Students would attend school normally in their designated classroom during the study.

### Measurements

The same measurements were conducted at baseline and at the 1-year visit. The measurements included ocular health, visual acuity, cycloplegic autorefraction and ocular biometry. The examination was conducted by the same group of 1 certified ophthalmologist, 3 certified optometrists and 2 certified ophthalmic nurses. Ocular health was examined using slit-lamp and direct ophthalmoscopy examinations. The Hirschberg test and cover test were used to identify ocular misalignment. Intraocular pressure was measured using a non-contact tonometer. Visual acuity was assessed with tumbling E Snellen visual charts (66 vision Medical Apparatus, Suzhou, China). Autorefraction was performed after full cycloplegia using Topcon 8800K (Topcon, Japan).

Cycloplegia was produced with 2 drops of 1% cyclopentolate (Cyclogyl, Alcon, Geneva, Switzerland) following 1 drop of Alcaine (Proparacaine Hydrochloride 0.5%, Alcon, Geneva, Switzerland), and full cycloplegia was assumed if the pupil diameter was greater than 6mm without a light reflex. Ocular biometry was measured using IOL master (Carl Zeiss Meditec, Jena, Germany). Final values were recorded by averaging three valid measurements.

### Outcomes

The primary outcome was the change of cycloplegic spherical equivalent (SER, spherical power + 1/2 cylindrical power) from baseline at one year, where SER was defined as spherical power plus one half of cylindrical power. The secondary outcomes at one year were the change of axial length (AL), and the incidence of myopia. Myopia was defined as SER ≤-0.50D, emmetropia as -0.50D<SER<+1.00D, and hyperopia as SER≥+1.00D after cycloplegia, where SER was the average of both eyes. Incidence of myopia was the primary outcome in the original study design, however, due to its definition, it had limitations to reflect the change of refractive error in baseline hyperopic and baseline myopic subjects. Change of cycloplegic spherical equivalent, on the other hand, is continous in nature and can reflect changes in all subjects.

### Power and Sample size

Sample size was calculated by comparing proportions between two groups, using the following formula:

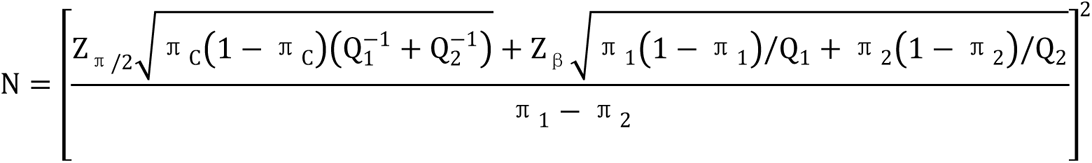

Based on a power of 80% (β=0.2), alpha of 0.05, the 2-year cumulative myopia incidences was expected 20% in the NC group, and 32 % in the TC gorup, the calculated sample size for each group was 206 patients. Allowing for participation rate of 90%, loss to follow-up of 5%, a final sample size of 243 per group would be used.

Based on current 1-year results, the mean difference of change of SER between the two groups was 0.17 D, with standard deviation of 0.62 D, by assuming 238 subjects in each group, alpha of 0.05, the corresponding power was 85%.

### Statistical Analysis

When analyzing SER and AL, eye-level data was used, while for other variables, patient-level data was used. Subgroup analyses were conducted by stratification of baseline refractive error status (hyperopia, emmetropia, and myopia). Patient-level continuous variables were summarized by means and standard deviations (SD), and comparisons between intervention and control group were carried out using T-test. For categorical variables, they were summarized by frequencies and proportions and compared between the intervention and control group utilizing Chi-squared test. Eye-level continuous variables (SER and AL) were summarized by model-based means and standard error (SE). The model used to compare SER and AL between intervention and control group was a generalized linear model using a Generalized Estimating Equation (GEE) approach.

Generalized linear model with GEE approach was generated to assess the risk factors associated with myopic shift (change of SER). Univariable analysis was first carried out in variables include treatment group, age, gender, baseline SER, and baseline AL. Risk factors with P-value less or equal to 0.2 were then put into multivariate model selection process. The final model was selected using backward selection. All analyses were conducted using SAS 9.4 (SAS Institute, Carry, NC, USA). Two-sided P<0.05 was considered statistically significant.

## Results

### Participants

A total of 483 students (955 eyes) from 10 classes agreed to participate this study, 5 classes were allocated randomly to the Outdoor Scene Classroom (OSC) group, and the other 5 classes were allocated randomly to the Traditional Classroom (TC) group. (**Figure 1**). At the one-year follow-up visit, 4 students were lost to follow-up and 33 did not complete the examinations (mainly due to refusing cycloplegia). As a result, 245 students in OSC group and 238 students in TC group were included in the final analysis. The retention rate was 93.2% in the OSC group, and 92.6% in the TC group.

**Figure 1.**
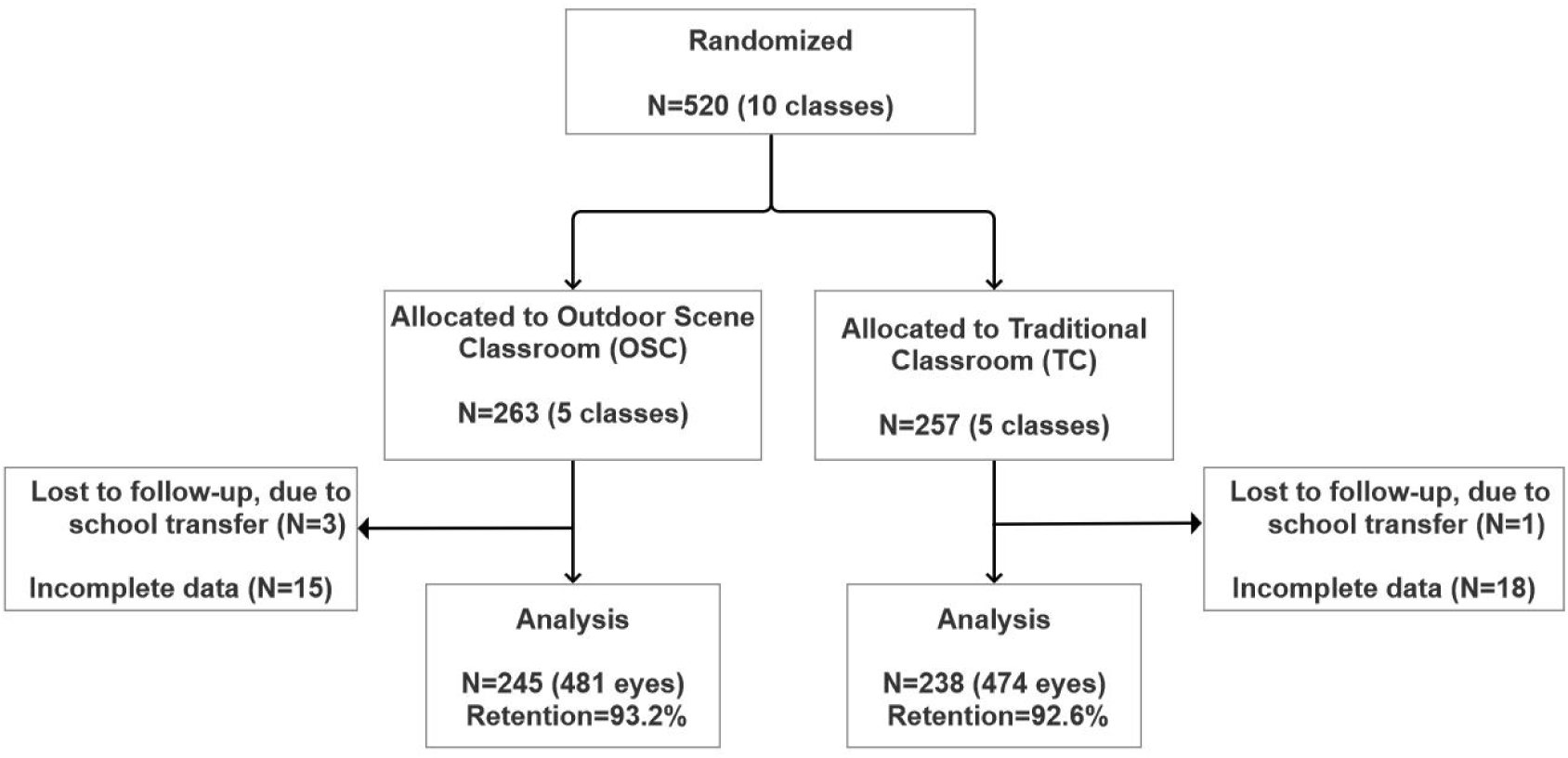
Flow chart.

### Baseline Characteristics

As shown in **Table 1**, the baseline characteristics of the OSC group and TC group were comparable, all no significant differences detected.

**Table 1:** Baseline characteristics of the participants.

| Characteristics | OSC<br>N=245 | TC<br>N=238 | P-value |
| --- | --- | --- | --- |
| Age in years, Mean (SD) | 9.26 (0.43) | 9.24 (0.48) | 0.63 |
| Gender |  |  | 0.76 |
| Male | 116<br>(47.3%) | 116<br>(48.7%) |  |
| Female | 129<br>(52.7%) | 122<br>(51.3%) |  |
| Spherical Equivalent in diopter, Mean (SE)* | 0.46 (0.09) | 0.58 (0.07) | 0.30 |
| Axial Length in mm, Mean (SE)* | 23.25<br>(0.06) | 23.14<br>(0.05) | 0.16 |
| Refractive error status |  |  | 0.53 |
| Myopia | 39 (15.9%) | 30 (12.6%) |  |
| Emmetropia | 122<br>(49.8%) | 119<br>(50.0%) |  |
| Hyperopia | 84 (34.3%) | 89 (37.4%) |  |

\*Generalized linear model-based results using GEE approach

**Table 2:** Change in refractive error,axial length and myopia incidence over one year.

| Characteristics | OSC<br>N=245 | TC<br>N=238 | P-value |
| --- | --- | --- | --- |
| Change of Spherical Equivalent in diopter, Mean (SE)* |  |  |  |
| All students | -0.49 (0.04) | -0.66 (0.03) | 0.002 |
| Hyperopic at baseline | -0.30 (0.04) | -0.52 (0.05) | 0.002 |
| Emmetropic at baseline | -0.48 (0.05) | -0.66 (0.05) | 0.01 |
| Myopic at baseline | -0.98 (0.13) | -1.05 (0.08) | 0.66 |
| Change of Axial Length in mm, Mean (SE)* |  |  |  |
| All students | 0.26 (0.01) | 0.31 (0.02) | 0.08 |
| Hyperopic at baseline | 0.17 (0.02) | 0.27 (0.05) | 0.04 |
| Emmetropic at baseline | 0.28 (0.02) | 0.29 (0.02) | 0.52 |
| Myopic at baseline | 0.43 (0.04) | 0.45 (0.03) | 0.65 |
| Myopia incidence, n (%) | 27 (13.1%) | 34 (16.3%) | 0.35 |

\*Generalized linear model-based results using GEE approach

**Table 3:** Risk factors associated with myopic shift.

|  | Univariable |  | Multivariate |  |
| --- | --- | --- | --- | --- |
| Characteristics | $\beta$ (95% CI) | P-value | $\beta$ (95% CI) | P-value |
| Group |  | 0.002 |  | <0.001 |
| OSC | Ref. |  | Ref. |  |
| TC | -0.16 (-0.26, -0.06) |  | -0.18 (-0.27, -0.09) |  |
| Age (Years) | -0.03 (-0.15, 0.09) | 0.61 | NA | NA |
| Gender |  | 0.61 |  | NA |
| Male | Ref. |  | NA |  |
| Female | 0.03 (-0.08, 0.13) |  | NA |  |
| Baseline SER (D) | 0.16 (0.10, 0.22) | <0.001 | 0.16 (0.10, 0.23) | <0.001 |
| Baseline AL (mm) | -0.15 (-0.24, -0.06) | 0.005 | NA | NA |

### Primary outcome

The overall change of SER was significantly different between the two groups. The mean (SE) change of SER was -0.49 (0.04) D in the OSC group and -0.66 (0.03) D in the TC group with P-value of 0.002. When stratified by student’s refractive error status as baseline, hyperopic and emmetropic strata showed significant differences between the OSC group and the TC group.

Among the baseline hyperopic students, the mean (SE) change of SER was -0.30 (0.04) D in OSC group and -0.52 (0.05) D in TC group (p=0.002). Among the baseline emmetropic students, the mean (SE) change in SER was -0.48 (0.05) D in OSC group and -0.66 (0.05) D in TC group (p=0.01). Among the baseline myopic students, however, the change of SER was not significantly different between the two groups.

### Secondary outcomes

The overall change of AL was not significantly different between the two groups. Among the baseline hyperopic students, the mean (SE) change of AL was 0.17 (0.02) mm in OSC group, significantly smaller than 0.27 (0.05) mm in TC group (p=0.04). Both the baseline emmetropic and the baseline myopic strata, showed no differences in change of AL between OSC group and TC group.

There were 206 non-myopic students in OSC group at baseline, 27 of them turned myopia by the end of year one with 13.1% of incidence rate. In the TC group, 34 out of 208 (16.3%) baseline non-myopic students became myopic. The difference was not statically significant (p=0.35).

### Safety

There were no adverse events related to the outdoor scene classroom intervention.

### Compare to studies of time outdoors and low-dose atropine

The effect of outdoor scene classroom (OSC) on children’s myopic shift were compared to time outdoors (Figure 2a) and low-dose atropine (Figure 2b). In comparing to the time outdoors, among non-myopic children, the myopic shift in OSC group was 0.19D less than the TC group, with 31.7% efficacy. The two-year cumulative effect of time outdoors from Shanghai Time Outside to Reduce Myopia trial (STORM) [12] was 0.20D less than the control, with 19.2% efficacy. The three-year cumulative effect of time outdoors from Guangzhou Outdoor Activity Longitudinal Study (GOALS) [13] was 0.17D less than the control, with 10.7% efficacy.

**Figure 2:**
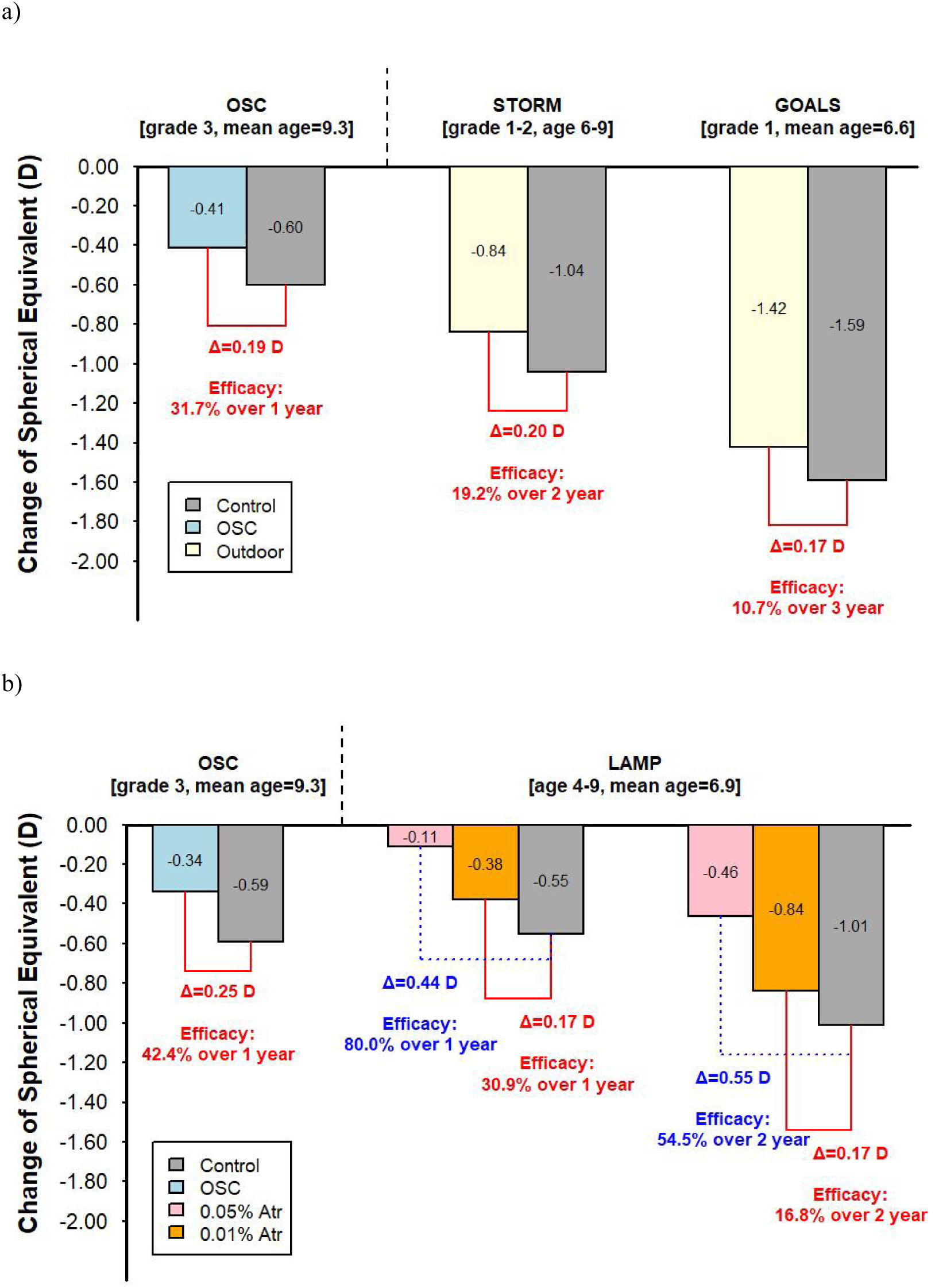
Demonstration of myopia prevention effect between the present study testing Outdoor Scene Classroom (OSC) and previous studies testing a) Time Outdoors and b) Low-dose Atropine. To keep the consistency of baseline spherical equivalent between the studies, only participants with baseline spherical equivalent above -0.50D from the present study was included in Panel a), and only with baseline spherical equivalent 0.00D<SER≤+1.00D was included in Panel b). STORM: Shanghai Time Outside to Reduce Myopia trial [10]; GOALS: Guangzhou Outdoor Activity Longitudinal Study [11]; LAMP: Low-Concentration Atropine for Myopia Progression trial [12].

In comparing to the low-dose atropine, among children with baseline SER between 0.00D and +1.00D, the myopic shift in OSC group was 0.25D less than the TC group, with 42.4% efficacy. The one-year effect of 0.01% and 0.05% atropine from Low-Concentration Atropine for Myopia Progression (LAMP) [14] was 0.17D, 0.44D less than the control, with 30.9%, 80.0% efficacy, respectively. The two-year cumulative effect was 0.17D, 0.55D less than the control, with 16.8%, 54.5% efficacy, respectively.

### Associated risk factors of one-year myopic shift

In the univariable models, the annual change of SER in the TC group is -0.16 (95% CI: -0.26, -0.06) D than it in the OSC group (p=0.002). Every 1 D increase in baseline SER is associated with 0.16 (95% CI: 0.10, 0.22) D less myopic shift (p<0.001). Every 1 mm increase in baseline AL is associated with -0.15 (95% CI: -0.24, -0.06) D more myopic shift (p=0.005). Age and gender were not significantly associated with myopic shift. In the multivariate final model, group (β =-0.18, p<0.001) and baseline SER (β=0.16, p<0.001) were remained.

## Discussion

In this randomized controlled trial, the outdoor scene classroom (OSC) showed protective effect in slowing down myopic shift (or preserving hyperopia reserve) in one year among children who were not myopic at baseline. The one-year SER change showed 0.22D less among baseline hyperopic children, and 0.18D less among baseline emmetropic children in the OSC group compare to TC group. It seems the protective effect of OSC is comparable to the effect of time outdoors and the 0.01% atropine, but lower than the 0.05% atropine. The absolute and relative myopia control effect in one year of OSC was 0.19D and 31.7% among non-myopic children at baseline, 0.25D and 42.4% among children with SER between 0.00D and +1.00D at baseline.

Similar effects were achieved with time outdoor studies of STORM (0.20D, 19.2%), GOALS (0.17D, 10.7%), and the 0.01% atropine from LAMP (0.17D, 30.9% in one year; 0.17D, 16.8% in two year). However, the effect was lower than the 0.05% atropine from LAMP (0.44D, 80% in one year; 0.55D, 54.5% in two year) [12–14].

The protective effect of OSC was strongest in the hyperopic children, whereas no difference was observed between the OSC group and the TC group in myopic children. In time outdoor studies, the same phenomenon has been observed. Consistent with our observations, in the STORM study, the authors reported time spent outdoors had no effect in myopia progression among myopic children [12]. The reason may be that myopic retina had different response to visual cues compared to emmetropic and hyperopic eyes. Swiatczak and Schaeffel have suggested myopic eyes may have reduced ability to detect myopic defocus [15–17].

These findings carry substantial public health implications, especially given the concerns around early-onset myopia, which has the risk to evolve into high myopia and severe ocular diseases [18]. Preventing the onset of myopia is in principle the best strategy to avoid the complications associated with high myopia. Education and reduced outdoor time have been identified as two major risk factors for myopia [19]. In Taiwan, the implementation of daily outdoor activities for two hours has been shown to decrease the prevalence of myopia among schoolchildren [20].

However, the applicability of a similar approach on a nationwide scale in mainland China raises questions. For example, He et al. highlighted the challenges of ensuring more than 80 minutes of outdoor activity during school days in Shanghai as part of the STORM study, indicating low feasibility [12].

In this context, the outdoor scene classroom offers an alternative strategy to increased time outdoors. Children are exposed to the OSC stimulus for all the time that they are in their classroom, approximately 7 hours per day, which provides significant protection. It may be difficult to increase this duration of exposure, but it may be possible to increase the efficacy of this approach by altering the spatial frequency content to the classrooms. More importantly, this method is a passive intervention; it does not demand extra effort from students, who simply attend their usual classes within an OSC setting. This strategy is not only cost-effective but also simple to implement, making it an attractive option for widespread adoption. Additionally, the design of the outdoor scene classroom has been well-received by both students and teachers [10], further supporting its potential for widespread adoption in efforts to combat myopia in children.

This approach could also be combined with increased outdoor exposure to prevent childhood myopia. Since current duration of exposures to time outdoors are limited, most commonly aiming for about 2 hours/day, it may be possible to increase these exposures somewhat, and if an additive effect is observed. The promising aspect is that the “Double Reductions Policy” implemented in China has the potential to reduce educational demands during preschool and early primary school years, which may result in more time spent outdoors [21].

The mechanism involved is not clear, but a possible hypothesis is that conditions that lead to the development of myopia lead to reductions in retinal dopamine release, whereas exposure to conditions that increase dopamine release arrest the development of myopia, as does treatment with dopamine agonists. This has been well-documented for the FDM and LIM experimental animal models [23–25], and is consistent with the hypothesis that more time outdoors will lead to more dopamine release [26]. While this mechanism needs to be further explored and tested, it provides a simple hypothesis for linking these two approaches to myopia prevention.

## Limitations

There were several limitations in this study need to be addressed. Firstly, the trial is not masked to the participants due to the nature of the intervention. Secondly, there was only one type of natural scene tested in this study. It’s worthwhile to investigate the threshold of myopia prevention effect of different spatial profiles in the future. Thirdly, the trial was initially designed as a two-year study. However, after one year the trial had to be terminated given the fact that students were re-allocated to a different campus due to school administrative reasons. Fortunately, we did find protective effect of OSC on arresting myopia development already at this timepoint for those students who were hyperopic at baseline. In addition, our results indicate that the protective effect of OSC attenuate with time, when refractive error naturally shifts from hyperopia to emmetropia with increasing age. Thus, the grade 3 students may not be the best age to observe the myopia prevention effect. Further study should be tested in a younger cohort where hyperopia is more prevalent (eg., grade 2, grade 1 or even preschool), and should be observed for longer term.

## Conclusion

In conclusion, the study found that the outdoor scene classroom was effective in reducing myopic shift and axial length elongation in hyperopic students compared to those in a traditional classroom setting. Large implementation of outdoor scene classroom provides an alternative strategy to increased time outdoors in myopia prevention, and provides an approach that involves less disruption to school routines. To further validate these findings, future studies should consider including a younger cohort of participants and exploring the impact of varying spatial frequency profiles, as well as combination with school routines that also increase time outdoors.

## Acknowledgement

We would like to thank Lun Pan in assisting the initial trial design and the process of randomization. All named authors meet the International Committee of Medical Journal Editors (ICMJE) criteria for authorship for this manuscript, take responsibility for the integrity of the work, and have given final approval to the version to be published.

## Author Contributions

Idea conception: W.L, D.I.F

Study design: W.P, W.L, Z.Y

Data preparation: L.W, X.Y, Y.G, Y.Z, Z.L, X.L

Data analysis: W.P, W.L

Manuscript drafting: W.P, W.L

Manuscript criticizing and revising: I.M, D.I.F, W.L, Z.Y

## Ethics statement

The study adhered to the Declaration of Helsinki and had been approved by Human Subjects Ethics Committee of the Aier School of Ophthalmology, Central South University (AIER2021IRB02). The trial was registered in the Chinese Clinical Trial Registry (https://www.chictr.org.cn; Registration Number: ChiCTR2000040704). Written informed consent were signed by both students and their guardians.

## Data Availability Statement

The data that support the findings of this study are not openly available due to reasons of confidentiality. Upon reasonable request, deidentified data can be accessed. Please contact corresponding author, Prof. Weizhong Lan.

## Code Availability Statement

Code can make available upon reasonable request, the code generated in the study was statistical codes using software SAS 9.4 (SAS Institute, Carry, NC, USA). Please contact corresponding author, Prof. Weizhong Lan.

